# Survival analysis of all critically ill patients with COVID-19 admitted to the main hospital in Mogadishu, Somalia, 30 March–12 June 2020: what interventions are proving effective?

**DOI:** 10.1101/2021.01.01.20248966

**Authors:** Mohamed M. Ali, Sk Md Mamunur Rahman Malik, Abdulrazaq Yusuf Ahmed, Ahmed Mohamed Bashir, Abdulmunim Mohamed, Abdulkadir Abdi, Majdouline Obtel

## Abstract

**OBJECTIVES:** To determine risk factors for death in patients with COVID-19 admitted to the main public sector hospital in Somalia and identify interventions contributing to improved clinical outcome in a low-resource and fragile setting.

**SETTING:** Main public sector tertiary hospital in Mogadishu, Somalia.

**PARTICIPANTS:** All 131 laboratory-confirmed COVID-19 patients admitted to the main public tertiary hospital in Somalia between 30 March and 12 June 2020.

**MAIN OUTCOME MEASURES:** We extracted demographic and clinical data from hospital records of all 131 COVID-19 patients admitted to hospital until their death or recovery. We used Kaplan–Meier statistics to estimate survival probabilities and the log-rank test to assess significant differences in survival between groups. We used the Cox proportional hazard model to estimate likelihood of death and assess the effect of risk factors on survival.

**RESULTS:** Of the 131 patients, 52 (40%) died in the hospital and 79 (60%) survived to discharge. The factors independently associated with increased risk of in-hospital death were: age ≥ 60 years – survival probability on day 21 in patients < 60 years was 0.789 (95% confidence interval (CI): 0.658–0.874) compared with 0.339 (95% CI: 0.205–0.478) in patients ≥ 60 years; cardiovascular disease (survival probability 0.478 (95% CI: 0.332–0.610) in patients with cardiovascular disease compared with 0.719 (95% CI: 0.601–0.807) in patients without cardiovascular disease); and non-invasive ventilation on admission – patients who were not ventilated were significantly more likely to survive than those who were (*P* < 0.001).

**CONCLUSION:** Our study, which includes the largest cohort of COVID-19 patients admitted to a single hospital in a sub-Saharan African country, confirms that underlying conditions and age are associated with increased risk of in-hospital death in patients with COVID-19. Our results show the advantage of medical oxygen over non-invasive ventilation in the treatment of patients with severe COVID-19 symptoms.

## Background

On 16 March 2020, the Federal Ministry of Health and Human Services of Somalia reported the country’s first laboratory-confirmed case of coronavirus disease-19 (COVID-19) in a Somali student arriving from China [1]. Since then and until 15 September 2020, the country has reported 3412 laboratory-confirmed cases of COVID-19, including 98 associated deaths out of 24 320 samples tested [2]. Of these cases, 639 cases (19%) with COVID-19 were admitted to 19 isolation centres designated by the government that were set up across the country. Of the cases hospitalized, only 242 patients presented with severe symptoms that needed critical care support in a tertiary-level hospital.

Given the fragility of the health care system in Somalia, it was anticipated that the system would be overwhelmed. The outbreak occurred at a time when the country had no intensive care beds, ventilators or a central supply of medical oxygen in the public sector. Ranked 193 out of 195 countries on the Global Health Security Index [3,4], Somalia’s health system has been weakened by decades of civil war, insecurity and disease outbreaks, as well as natural disasters such as droughts and floods. The current health workforce density in Somalia (0.34 health care workers per 1000 population) is substantially lower than the density needed for universal health coverage (UHC) – 4.45 health care workers per 1000 population by 2030 [5,6]. At the time the epidemic hit the country, there were only 15 intensive care unit beds (all in the private sector) for a population of more than 15 million. The disease surveillance system to detect, monitor and facilitate a rapid response to any outbreak is dysfunctional and fragmented.

To date, most of the laboratory-confirmed cases of COVID-19 in Somalia did not require hospitalization as over 80% were either asymptomatic or had mild symptoms (unpublished data from case-based surveillance, 2020). Between 16 March and 16 September 2020, out of 13 800 people who were tested in Mogadishu, 1531 (11%) were confirmed with COVID-19, 55 of whom died. Of these 1531 cases, only 131 with severe clinical disease were admitted to the main hospital of the country in Mogadishu.

Although data are available from developed and high-income countries on the clinical characteristics of COVID-19, and the outcome and risk factors for clinical outcome [5]8], few studies investigating the links between interventions and clinical outcome have been published from less developed countries [6].

Documenting the length of hospitalization and survival of patients with COVID-19 and the risk factors associated with death in low-resource settings could provide a better understanding of the impact of the disease and the usefulness of medical interventions as well as hospital capacity to cope with the surge in COVID-19 patients in such settings. This information can also guide policy responses on the use of low-cost, high-impact interventions in such setting to save lives and manage a surge in cases.

The main aim of this study was to assess the clinical characteristics of patients with COVID-19 admitted to the main public-sector hospital in Somalia, estimate the length of hospital stay and identify the risk factors for death in these patients. The study also aimed to consider what interventions might help improve clinical outcomes in patients with severe COVID-19 in low-resource and fragile settings.

## Methods

### Data extraction and outcomes

We extracted information from clinical records on all 131 COVID19 patients admitted to De-Martino hospital, Mogadishu between 30 March 2020 and 12 June 2020. The information extracted included: patient’s place of residence, age, sex, date of admission, date of onset of symptoms, reported signs and symptoms (fever, shortness of breath, muscular pain, chest pain, abdominal pain, joint pain, general weakness, diarrhoea, cough, nausea or vomiting, sore throat, headache, runny nose, irritability and confusion), clinical characteristics (temperature, pharyngeal exudate, coma, abnormal lung X-ray, conjunctival infection, dyspnoea or tachypnoea, seizure and abnormal lung auscultation), comorbidities as reported by the patients (cardiovascular disease including hypertension, immunodeficiency including HIV, diabetes, renal disease and chronic lung disease), clinical interventions (delivery of oxygen (ventilation support through a facemask under positive pressure without the need for endotracheal intubation, non-invasive ventilation, inotropes and vasopressors, and antibiotics), and final outcome and date (survived or died). We transcribed individual patient data into an *Excel* file, and both the care providers and the information officers independently verified the data from the patient medical records for completeness and accuracy.

The primary outcome was survival status and time to outcome from date of hospitalization (in days).

### Statistical analysis

We report the distribution of cases and deaths by sex and age group, and the distribution of symptoms, clinical characteristics, comorbidities and type of treatment by sex and age group. We used Kaplan–Meier statistics to estimate survival probabilities and the log-rank test to assess the significance of differences in survival between groups. We extended the survival analysis using the Cox proportional hazard model to estimate the likelihood of death and simultaneously assess the effect of several risk factors on survival, adjusting for confounding or effect modification. We also examined the assumption of proportionality.

We report numbers and percentages for binary variables, mean and median for continuous variables. We dichotomized age into < 60 years and ≥ 60 years. We used Stata, version 16 for data quality assessment and analysis. *P* < 0.05 was considered statistically significant.

### Patient and public involvement

No patient involved

## Results

We followed all 131 patients with COVID-19 admitted to the hospital until the end of their stay. The first admission was on 30 March 2020 and the last was on 12 June 2020. The final date of discharge or death was on 15 June 2020. The length of hospital stay ranged between 1 day and 35 days with a median length of 5.5 days, interquartile range (IQR) 9 days.

Two thirds (89/131; 68%) of the patients were males, with mean age of 58.5 (SD: 16.1) years and median of 60 (IQR: 18) years) for males and 56.9 (SD: 17.1) years and median 55 (IQR: 27) years for females. Of the 131 patients, 52 (40%) died (Table 1).

**Table 1:** Number of patients with COVID-19 and deaths associated with COVOD-19, by age and sex, Somalia, 2020.

| Age group (years) | Males |  | Females |  |
| --- | --- | --- | --- | --- |
|  | Total no. of patients | No. died | Total no. of patients | No. died |
| 10–19 | 2 | 0 | 0 | 0 |
| 20–29 | 3 | 0 | 4 | 2 |
| 30–39 | 9 | 0 | 6 | 0 |
| 40–49 | 5 | 2 | 4 | 0 |
| 50–59 | 15 | 4 | 11 | 4 |
| 60–69 | 32 | 19 | 9 | 4 |
| 70–79 | 17 | 8 | 3 | 1 |
| 80+ | 6 | 5 | 5 | 3 |
| **Total** | **89** | **38** | **42** | **14** |

Table 2 shows the distribution of patients according to signs and symptoms, clinical characteristics, comorbidities and the interventions given, by age group and sex. Most of the patients reported fever, shortness of breath and cough; few had abdominal pain, diarrhoea or a runny nose. Muscular and joint pain, general weakness and headache increased with age for both sexes. None of the patients presented with pharyngeal exudate, coma, conjunctival infection or seizures. However, 125/131 (95%) of patients had abnormal lung findings on X-ray and 124/131 (95%) abnormal lung auscultation, and 119/131 (91%) presented with dyspnoea and/or tachypnoea. More males had these clinical characteristics than females, but the frequency of the symptoms increased with age in females.

**Table 2:** Distribution of the patients according to disease characteristics and treatment, by sex and age group, Somalia, 2020.

| Variable | Females (no.) |  |  | Males (no.) |  |  |
| --- | --- | --- | --- | --- | --- | --- |
|  | Age (years) |  |  | Age (years) |  |  |
|  | < 40 | 40–59 | ≥ 60 | < 40 | 40–59 | ≥ 60 |
| **Symptoms and signs** |  |  |  |  |  |  |
| Fever | 9 | 13 | 17 | 13 | 19 | 54 |
| Shortness of breath | 8 | 15 | 16 | 14 | 20 | 52 |
| Muscular pain | 1 | 1 | 6 | 1 | 2 | 11 |
| Chest pain | 3 | 4 | 9 | 4 | 11 | 21 |
| Abdominal pain | 2 | 2 | 2 | 0 | 1 | 5 |
| Joint pain | 1 | 1 | 9 | 2 | 3 | 18 |
| General weakness | 3 | 4 | 10 | 3 | 6 | 24 |
| Diarrhoea | 1 | 1 | 1 | 0 | 2 | 5 |
| Cough | 9 | 15 | 16 | 14 | 19 | 52 |
| Nausea/Vomiting | 2 | 3 | 0 | 0 | 1 | 6 |
| Sore throat | 4 | 6 | 8 | 5 | 9 | 26 |
| Headache | 4 | 5 | 12 | 7 | 12 | 30 |
| Runny nose | 0 | 0 | 0 | 0 | 0 | 4 |
| Irritability confusion | 2 | 1 | 6 | 1 | 6 | 15 |
| **Clinical characteristics** |  |  |  |  |  |  |
| Abnormal lung X-ray findings | 7 | 13 | 17 | 14 | 20 | 54 |
| Dyspnea tachypnea | 8 | 14 | 17 | 14 | 19 | 47 |
| Abnormal lung auscultation | 7 | 14 | 17 | 14 | 20 | 52 |
| **Comorbidities** |  |  |  |  |  |  |
| Cardiovascular disease | 0 | 6 | 8 | 0 | 10 | 29 |
| Immunodeficiency, including HIV | 0 | 0 | 0 | 1 | 0 | 1 |
| Diabetes | 1 | 9 | 10 | 2 | 9 | 21 |
| Renal disease | 1 | 1 | 1 | 0 | 1 | 2 |
| Chronic lung disease | 1 | 1 | 2 | 0 | 1 | 10 |
| **Treatment** |  |  |  |  |  |  |
| Oxygen therapy | 7 | 14 | 17 | 11 | 17 | 53 |
| Non-invasive ventilation | 1 | 1 | 5 | 0 | 3 | 20 |
| Inotropes and vasopressors | 0 | 1 | 3 | 0 | 4 | 20 |
| Antibiotics | 10 | 15 | 17 | 14 | 20 | 55 |
| **Total** | **10** | **15** | **17** | **14** | **20** | **55** |

The main comorbidities were cardiovascular disease in 53/131 (40%) patients and diabetes in 52/131 (40%). Cardiovascular disease was reported only by patients 40 years or older. Diabetes was more prevalent in females, 20/42 (48%), compared with in males, 32/89 (36%). Only two male patients had immunodeficiency. None of the female patients was pregnant or reported end of pregnancy in the 6 weeks before hospitalization. All the patients were given antibiotics, and oxygen therapy was given to 119/131 (91%) patients; the use of oxygen increased with patient age. Inotropes and vasopressors were given to 28/131 (21%) patients, all of whom were 40 years or older.

Table 3 shows the survival probabilities with 95% confidence intervals (CI) at days 7, 14 and 21 after admission, by age group, sex, presence of cardiovascular disease and diabetes, and treatment with inotropes and vasopressors. Patients 60 years and older were significantly more likely to die than patients younger than 60 years (*P* < 0.003), and the risk of death was significantly different between the two age groups at days 7, 14 and 21 (Figure 1a). The probability of surviving on day 21 in patients younger than 60 years was 0.789 (95% CI: 0.658– 0.874) compared with 0.339 (95% CI: 0.205–0.478) in patients 60 years and older. The probability of death was very similar for males and females; *P* = 0.049 (Figure 1a).

**Table 3:** Survival probability according to patient characteristics and treatment on day 7, 14 and 21 after admission to hospital, Somalia, 2020.

| Variable | At day 7<br>Survival probability<br>(95% CI) | At day 14<br>Survival probability<br>(95% CI) | At day 21<br>Survival probability<br>(95% CI) |
| --- | --- | --- | --- |
| **Age group (years)** |  |  |  |
| < 60 | **0.789 (0.658–0.874)** | **0.789 (0.658–0.874)** | **0.789 (0.658–0.874)** |
| ≥ 60 | **0.489 (0.365–0.602)** | **0.440 (0.312–0.561)** | **0.339 (0.205–0.478)** |
| **Sex** |  |  |  |
| Female | 0.669 (0.497–0.794) | 0.669 (0.497–0.794) | 0.535 (0.254–0.752) |
| Male | 0.600 (0.487–0.696) | 0.557 (0.436–0.662) | 0.488 (0.348–0.613) |
| **Cardiovascular disease** |  |  |  |
| No | 0.719 (0.601–0.807) | 0.697 (0.574–0.790) | 0.558 (0.379–0.704) |
| Yes | 0.478 (0.332–0.610) | 0.425 (0.265–0.576) | 0.425 (0.265–0.576) |
| **Diabetes** |  |  |  |
| No | 0.704 (0.585–0.795) | 0.684 (0.561–0.779) | 0.547 (0.372–0.693) |
| Yes | 0.499 (0.353–0.629) | 0.449 (0.290–0.596) | 0.449 (0.290–0.596) |
| **Inotropes or vasopressors** |  |  |  |
| No | **0.769 (0.670–0.842)** | **0.728 (0.614–0.813)** | **0.655 (0.508–0.768)** |
| Yes | **0.107 (0.027–0.251)** | **0.107 (0.027–0.251)** | **0.000 (0.000–0.000)** |

CI: confidence interval.

**Figure 1:**
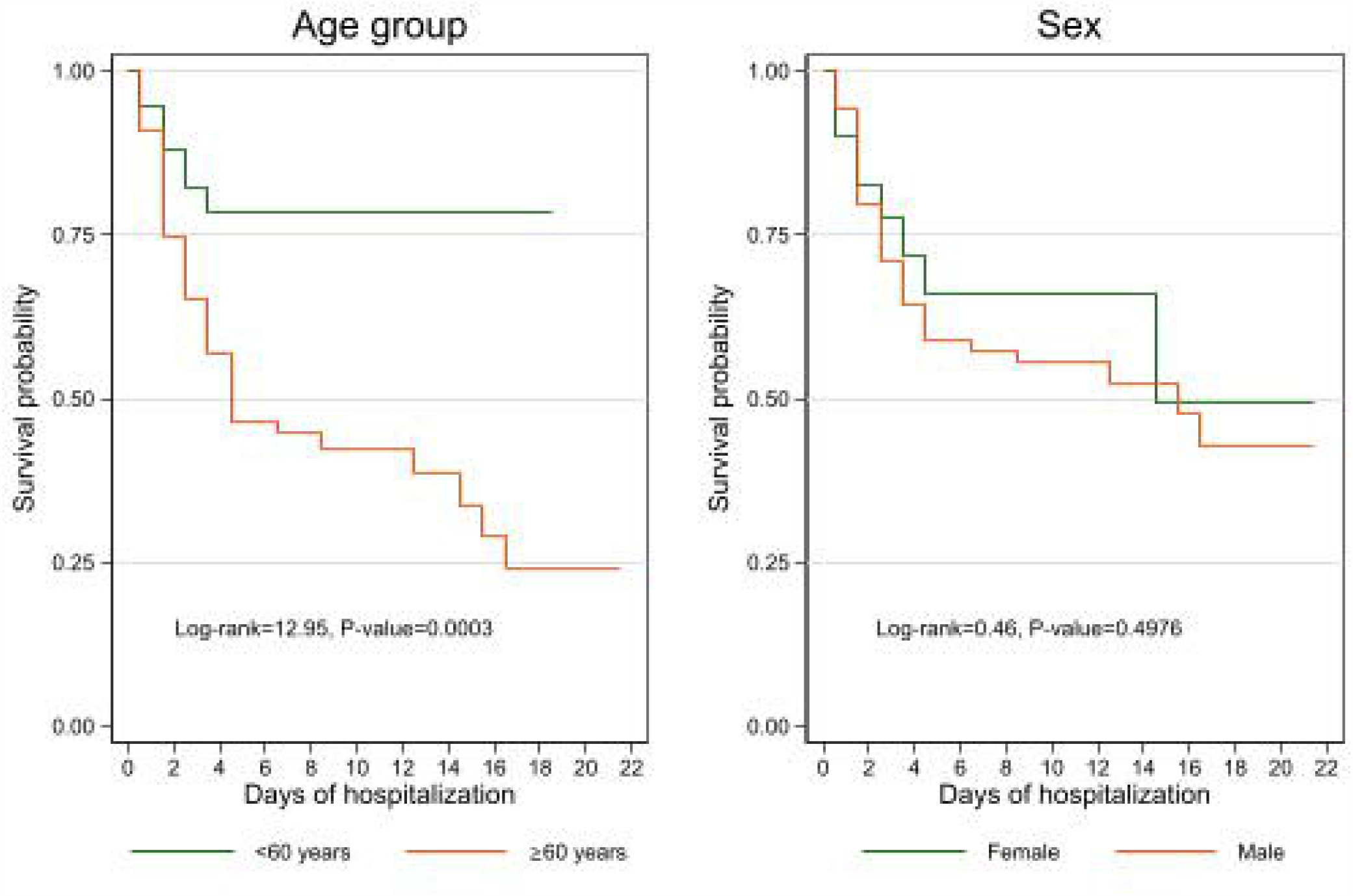
Kaplan-Meier survival estimates by age and sex.

Patients without cardiovascular disease were significantly more likely to survive than those with cardiovascular disease (*P* = 0.0134) (Figure 2). About half of the patients with cardiovascular disease survived the first week of hospitalization (survival probability = 0.478, 95% CI: 0.332– 0.610) compared with three-quarters of patients without cardiovascular disease (survival probability = 0.719, 95% CI: 0.601–0.807). However, the survival probabilities of patients without cardiovascular disease decreased by the third week and reached 0.558 (95% CI: 0.379– 0.704) compared with 0.425 (95% CI: (0.265–0.576) Table 3). A similar pattern was seen in patients with diabetes, although the difference was smaller (Figure 2).

**Figure 2:**
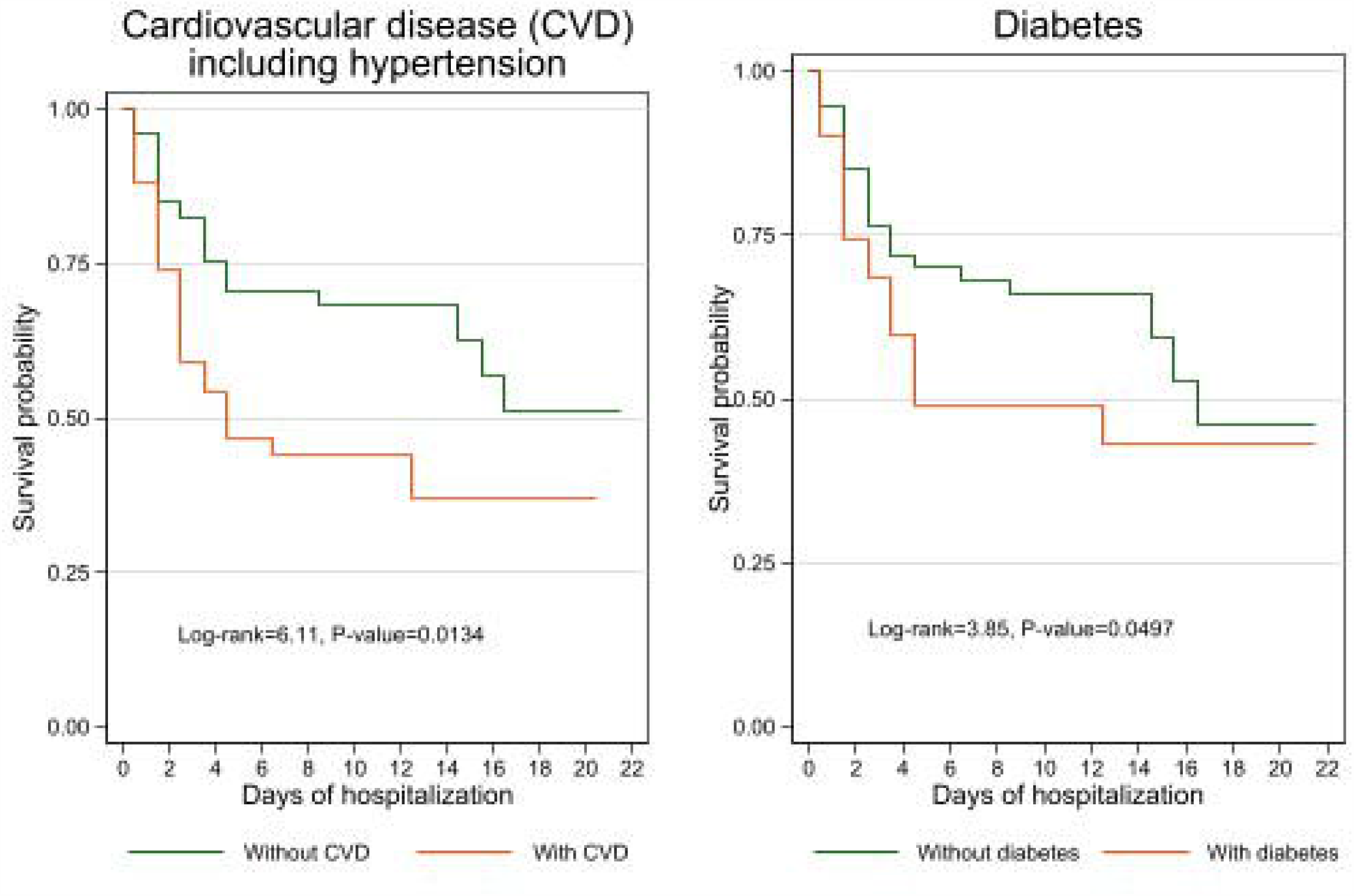
Kaplan-Meier survival estimates by comorbidity.

Of 29 patients given inotropes or vasopressors, 25 (86%) died in the first week of admission. The survival probability of those who did not receive this intervention was significantly higher at day 7 (survival probability = 0.769, 95% CI: 0.670–0.842), at day 14 (survival probability = 0.728, 95% CI: 0.614–0.813) and at day 21 (survival probability = 0.655, 95% CI: 0.508–0.768) after admission compared with those who received the intervention (Table 3). Overall, patients not given inotropes or vasopressors were significantly more likely to survive than those given them (*P* < 0.001) (Figure 3).

**Figure 3:**
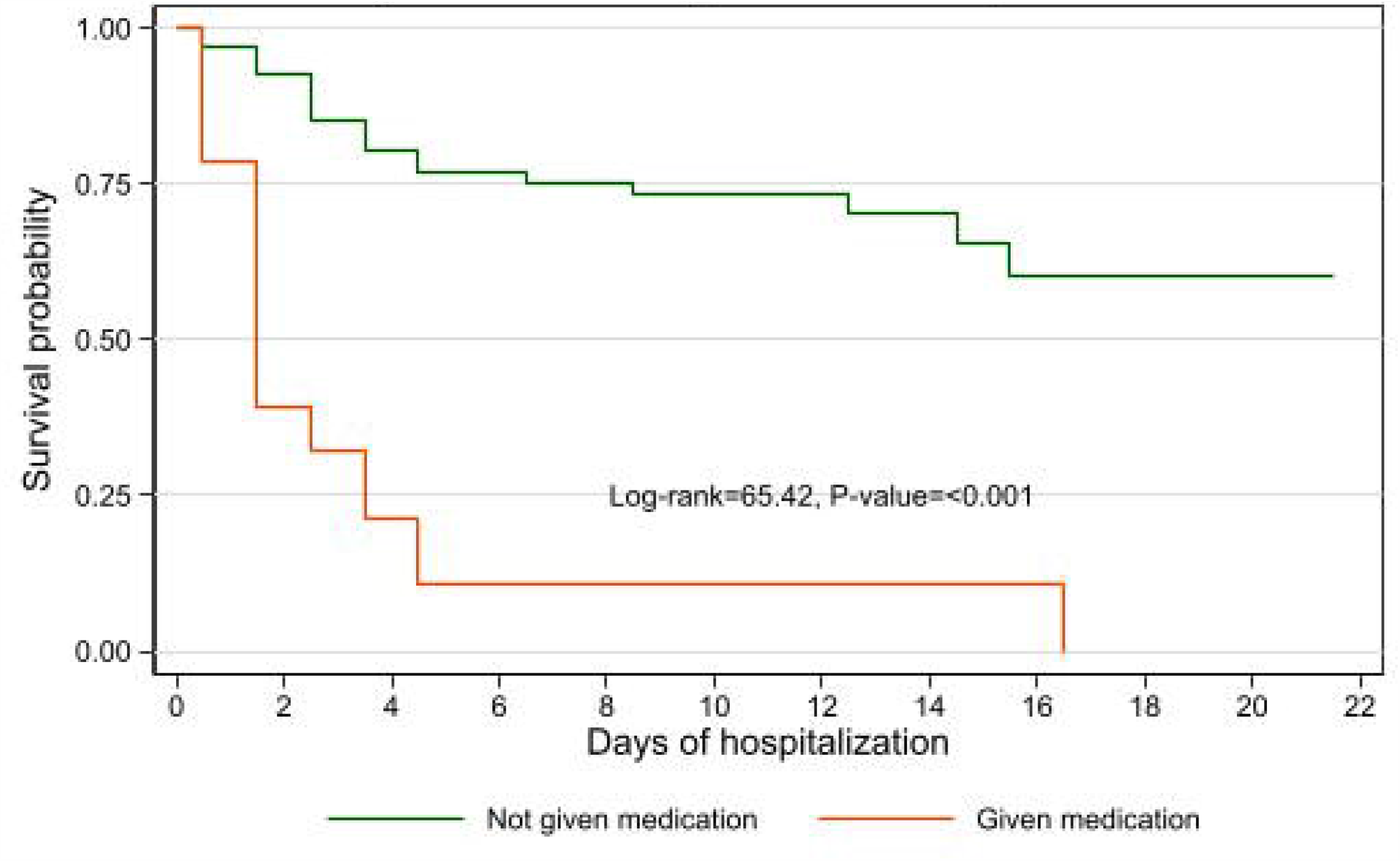
Kaplan-Meier survival estimates by use of inotropes and vasopressors.

Out of 131 patients, 12 were not given oxygen as their condition was considered mild and they all survived. We categorized the remaining 119 patients who were given oxygen on admission into three categories – not given non-invasive ventilation, delayed non-invasive ventilation and non-invasive ventilation given on admission – and calculated their survival probabilities (Figure 4 and Table 4). Those who were not ventilated or later ventilated had higher probability of survival than those who were ventilated on admission. Those who were ventilated on admission had a 22% chance of survival on day 3, compared with 83% on those who were not ventilated at all, and none of patients ventilated on admission survived after day 4 (Table 4). All the patients who were given delayed ventilation, when their health condition worsened, also died and did not survive beyond day 16. Overall, patients who were not ventilated were significantly more likely to survive than those who were ventilated (*P* < 0.001) (Figure 4).

**Table 4:** Survival probability according to length of time after admission, by administration of non-invasive ventilation, Somalia, 2020.

| Length of time after admission (days) | No ventilation | Delayed ventilation | Ventilation on admission |
| --- | --- | --- | --- |
|  | Survival probability (95% CI) | Survival probability (95% CI) | Survival probability (95% CI) |
| 1 | 0.966 (0.899–0.989) | 1.000 (1.000–1.000) | 0.739 (0.509–0.873) |
| 2 | 0.910 (0.827–0.954) | 1.000 (1.000–1.000) | 0.304 (0.135–0.493) |
| 3 | 0.828 (0.731–0.893) | 1.000 (1.000–1.000) | 0.217 (0.079–0.399) |
| 4 | 0.778 (0.675–0.853) | 1.000 (1.000–1.000) | 0.087 (0.015–0.242) |
| 5 | 0.753 (0.647–0.832) | 0.714 (0.258–0.920) |  |
| 7 | 0.753 (0.647–0.832) | 0.571 (0.172–0.837) |  |
| 9 | 0.737 (0.627–0.819) | 0.571 (0.172–0.837) |  |
| 14 | 0.737 (0.627–0.819) | 0.429 (0.098–0.734) |  |
| 15 | 0.737 (0.627–0.819) | 0.214 (0.012–0.586) |  |
| 16 | 0.698 (0.564–0.798) | 0.214 (0.012–0.586) |  |
| 21 | 0.698 (0.564–0.798) |  |  |

CI: confidence interval

**Figure 4:**
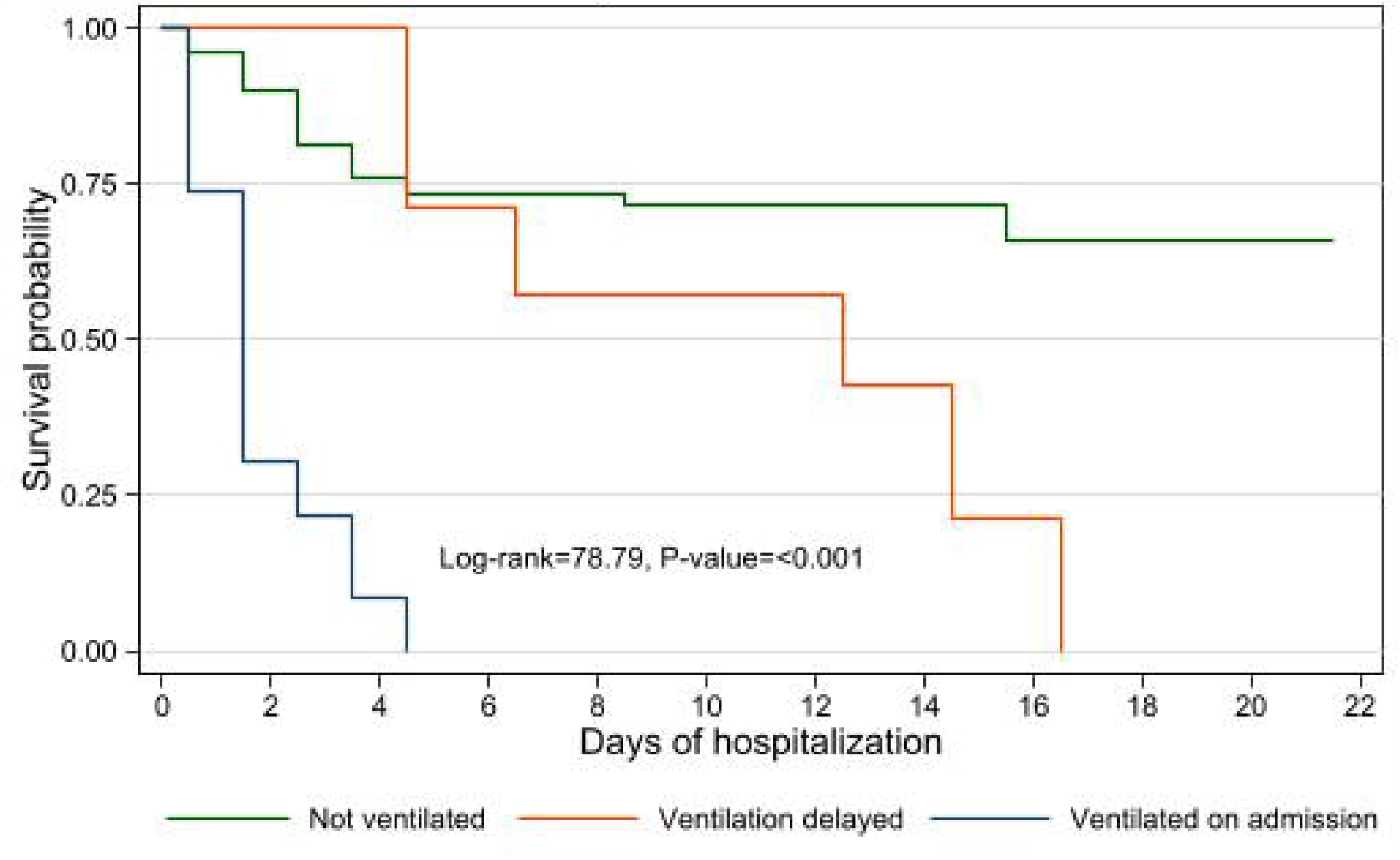
Kaplan-Meier survival estimates by ventilation use.

The likelihood of death for females and males by ventilation status was estimated using a Cox proportional hazard model, adjusting for age and two comorbidities. Table 5 shows the adjusted hazard ratios. The likelihood of death in female patients who were ventilated on admission (hazard ratio = 33.79, 95% CI: 4.09–279.07) was higher than in males who were ventilated on admission (hazard ratio = 8.65, 95% CI: 3.93–19.06).

**Table 5:**
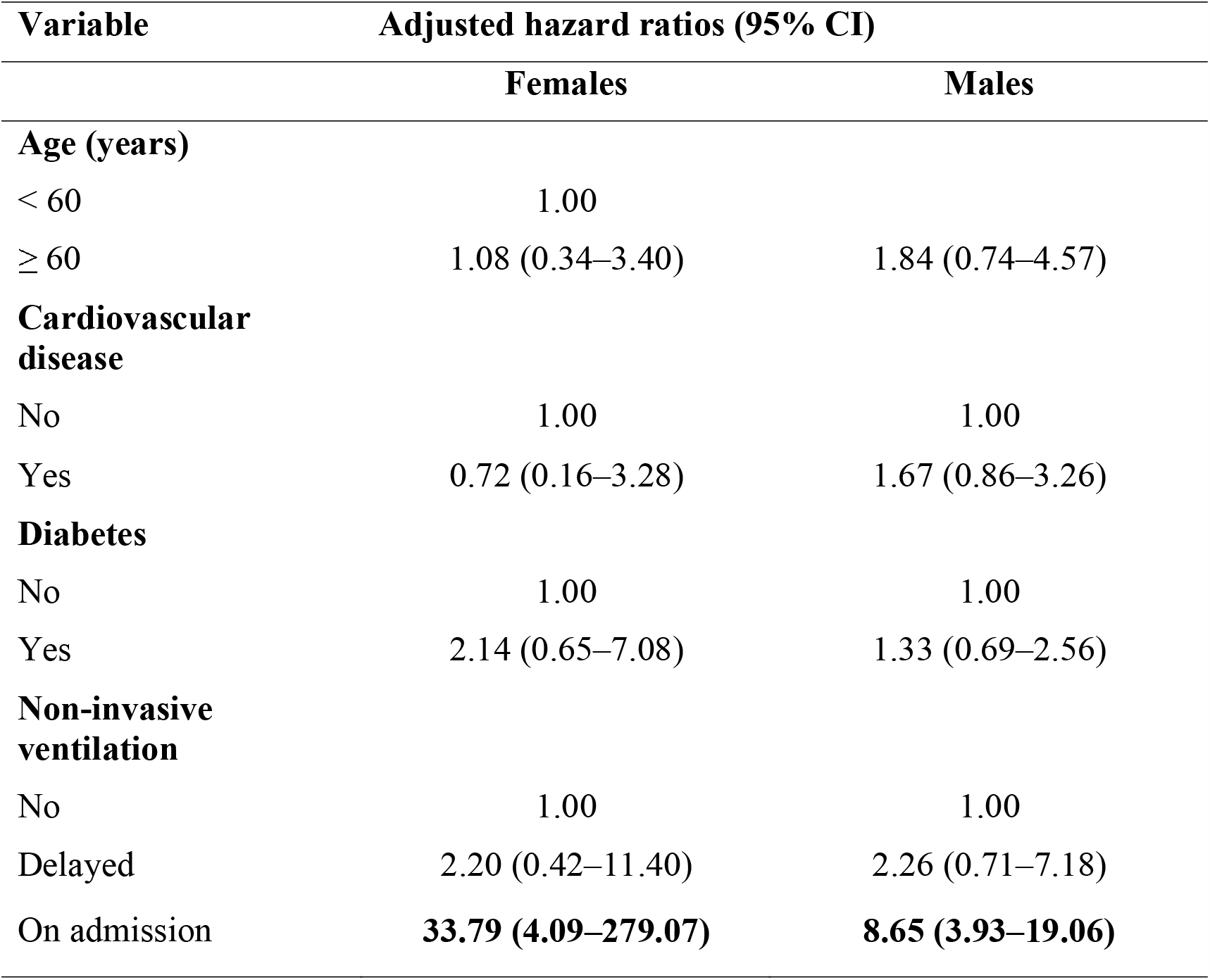
Adjusted hazard ratios for death in hospitalized patients with COVID-19, by sex, Somalia, 2020.

| Variable | Adjusted hazard ratios (95% CI) |  |
| --- | --- | --- |
|  | Females | Males |
| **Age (years)** |  |  |
| < 60 | 1.00 |  |
| ≥ 60 | 1.08 (0.34–3.40) | 1.84 (0.74–4.57) |
| **Cardiovascular disease** |  |  |
| No | 1.00 | 1.00 |
| Yes | 0.72 (0.16–3.28) | 1.67 (0.86–3.26) |
| **Diabetes** |  |  |
| No | 1.00 | 1.00 |
| Yes | 2.14 (0.65–7.08) | 1.33 (0.69–2.56) |
| **Non-invasive ventilation** |  |  |
| No | 1.00 | 1.00 |
| Delayed | 2.20 (0.42–11.40) | 2.26 (0.71–7.18) |
| On admission | **33.79 (4.09–279.07)** | **8.65 (3.93–19.06)** |

## Discussion

We assessed the demographic and clinical characteristics, outcomes and risk factors for death in all 131 patients with COVID-19 who were admitted to the De Martini hospital, the largest and only public sector hospital in Somalia, between 30 March and 12 June 2020. Before the COVID-19 outbreak, the hospital had no intensive care beds and it was not designed to care for severely ill patients. When COVID-19 began in the country, the government refurbished the De Martini hospital and fitted it with 20 intensive care beds. The decision was taken to transfer all severely ill patients diagnosed with COVID-19 to this hospital. However, there was no definitive criteria or strategy for discharging patients once admitted to the hospital.

Between 30 March and 12 June 2020, 79/131 patients (60%) were discharged alive and 52/131 (40%) patients died. To our knowledge, this study includes the largest cohort of patients with COVID-19 admitted to a single hospital in a sub-Saharan African country.

The median length of hospital stay was 5.5 days (IQR: 9 days) with a range of 1–35 days and a mean of 7.7 days (SD: 6.9). In a similar study in Vietnam, the median length of hospital stay was 21 (range: 16–34) days [6], while in the United States of America and several European countries, the length of stay was shorter with an average of 7–8 days only [10–12].

Two thirds (90/131; 69%) of the patients admitted in De Martini hospital during the study period were males, with mean age of 58.2 years for males and 56.9 years for females. These findings are consistent with other studies in Vietnam [6] and China [8] which suggest a slight gender difference in hospitalized patients with COVID-19. The signs and symptoms by age group and sex in our study do not differ from those reported in studies outside of Africa [8,9,14]; however there are no comparable data from African settings, especially in sub-Saharan African countries.

The main comorbidities found in the 131 patients with COVID-19 in our study are similar to the findings of other studies done in Africa [10] Although we are limited by the lack of population-based data on the prevalence of diabetes and cardiovascular diseases in males and females in Somalia, the fact that the proportion of female COVID-19 patients with diabetes in our study (48%; 20/42) was higher than males (32/89; 36%) is consistent with a study done in some areas of Somalia in 2016, which found that the prevalence of risk factors such as diabetes was higher in females than males, and CVD was higher among males, and in both sexes, the risk factors increased with age [15]. These findings are also consistent with another study that examined the burden of noncommunicable diseases in sub-Saharan Africa [12].

Our survival analysis and estimation of the hazard of death show that older age (> 60 years) significantly increased the likelihood of death due to COVID-19. However, patient’s sex did not increase the likelihood of death. In a study in China, men were more than twice as likely to die from COVID-19 than women, which differs from our study and age > 50 years increased the risk of death from COVID-19, which concurs with our study [13]. The use of vasopressors and inotropes did not reduce the risk of death, as patient received such treatment have very low probability of survival, and the adjusted hazard of death is 6.24 (95%CI: 3.55,10.99) compared to those who were not given the treatment (calculation not shown). Another study has reported similar results [8].

Similar to another study [18], we also found that the use of non-invasive ventilation was an indicator of severe disease and increased the risk of death. An important finding of our study is the comparative benefit in the clinical outcome of providing the patients with non-invasive ventilation and the use of medical oxygen. The survival probability of patients who were given medical oxygen only was higher (75%) at day 7, and consistently remained at over 70% even at day 14 after admission, than in patients treated with both oxygen and non-invasive ventilation (10%), the risk of death for patients given non-invasive ventilation with medical oxygen was 5.43 times higher than in patients given only oxygen (calculation not shown). The lack of clinical benefit from the use of ventilation has also been reported in other settings [11,19,[16].

As the COVID-19 pandemic spread around the world, there was concern that the infection would have a substantial impact on African countries because they were unprepared to deal with such a crisis [17][18]. While this is true, the level of preparedness was judged by the number and availability of intensive care beds and ventilators per million of people’s use. However, ventilators, although useful for patients suffering from severe symptoms, are not a feasible treatment in countries where skilled and trained staff, such as specialized nurses and intensivists to manage intensive care units and ventilators, are not available, as was the case in our hospital.

Our study has a number of limitations. First, the dataset does not include all clinical variables related to the evolution of COVID-19, and data were collected manually, although the data were checked by two people to minimize error. Nonetheless, human error and missing information should be taken into consideration while analysing and interpreting the data. Second, our data include information on deaths from laboratory-confirmed cases and do not include other patients who could not be tested. This may lead to underestimation of our findings (of the benefit of oxygen over non-invasive ventilation) and limit their generalizability.

Despite these limitations, our study contributes to an understanding of the risk factors for death from COVID-19 in Somalia. Risk factors for death from COVID-19 have not been studied or published from sub-Saharan countries before.

## Conclusion

To our knowledge, this is the first evidence from a fragile and resource-poor setting on the advantage of medical oxygen over non-invasive ventilation in the treatment and care of patients with severe symptoms of COVID-19. Information on the usefulness of medical oxygen is reported from the United States [15] and Italy [16], but there has been no such evidence from any African country so far. The study has important policy implications by highlighting the value of available, accessible and affordable low-cost interventions in a fragile and resource-poor setting.

As reported in most sub-Saharan African countries [19], medical oxygen in secondary and tertiary health care settings is not always available even though it is well established that this therapy is fundamental for the treatment of severely ill patients, especially for pneumonia, which is a leading cause of death in elderly people and children under 5 years. Its usefulness in the treatment of patients with COVID-19 is further evidence of the urgent need to ensure that medical oxygen is always available in these settings.

## Data Availability

the study data will be available from the WHO country office

## Acknowledgement

We would like to acknowledge Jonathan Aaron Polonsky for his review and Ms Fiona Curlet for her review and editing

## Patient consent for publication

Not required.

